# Evaluation of a flow cytometry-based surrogate assay (FlowSA) for the detection of SARS-CoV-2 in clinical samples

**DOI:** 10.1101/2024.06.12.24308675

**Authors:** Vinit Upasani, Marjolein Knoester, Daniele Pantano, Lili Gard, Jolanda M. Smit, Bernardina T.F. van der Gun, Adriana Tami, Izabela A. Rodenhuis-Zybert

## Abstract

The current diagnostic methods for SARS-CoV-2 rely on quantitative RT-PCR. However, the presence of viral RNA in samples does not necessarily reflect the presence of an infectious virus. Therefore, the reliable detection of infectious SARS-CoV-2 in clinical samples is necessary to limit viral transmission. Here, we developed a flow cytometry-based surrogate assay (FlowSA), wherein the presence of infectious SARS-CoV-2 was detected using virus nucleocapsid-specific antibodies. We showed that FlowSA allows the detection of a wide range of viral titers of multiple SARS-CoV-2 variants. Furthermore, the assay was successfully used to detect infectious SARS-CoV-2 in nasopharyngeal swabs from SARS-CoV-2 positive individuals, including those with high Ct values. Notably, FlowSA identified the presence of infectious SARS-CoV-2 in biological specimens that scored negative for cytopathic effect (CPE) in cell culture and would otherwise be considered negative. We propose that FlowSA can be adopted as an alternative to conventional CPE methods for viral diagnostics.

## Introduction

Coronavirus disease 2019 (COVID-19) is caused by the novel coronavirus, Severe Acute Respiratory Syndrome Coronavirus 2 (SARS-CoV-2) (1,2). The virus belongs to the subgenus Sarbecovirus, genus Betacoronavirus, and family *Coronaviridae*. Since the beginning of the pandemic, new variants of SARS-CoV-2 have emerged worldwide, namely Alpha (B.1.1.7), Beta (B.1.351), Gamma (P.1.), Delta (B.1.617.2) and the most recent lineage is Omicron (B.1.1.529). The variants have multiple substitutions in the surface spike protein, some of which are in the receptor-binding domain thereby influencing virus transmission (3).

Early detection of SARS-COV-2 infection is crucial to limit further transmission. Furthermore, once an infected individual has recovered, it is essential to determine whether the person is still actively shedding infectious virus. Currently, real-time PCR (RT-PCR) is the most frequently used method to detect viral RNA in nasopharyngeal specimens because of its reliability and sensitivity (4-6). However, one drawback of the RT-PCR method is its inability to discriminate between the total viral load and viable, i.e., infectious virus particles. Therefore, RT-PCR may have limited use as a tool to de-escalate and discontinue infection control or quarantine measures (7-9). For the detection of viable viruses from patient samples, cell culture-based methods, including a conventional plaque assay or 50% tissue culture infectious dose (TCID-50) assay, can be used. In these assays, the presence of viable virus particles in the samples is measured as a function of disruption of the cell monolayer, termed cytopathic effect (CPE), which can be observed visually. However, this technique has several constraints such as extended incubation times, limited specificity, and observer subjectivity. Moreover, when CPE is observed, additional validation using e.g. staining of viral proteins is required to determine the virus species (10-12).

Our objective was to develop a highly sensitive, virus-specific, and scalable method for high-throughput detection of infectious SARS-CoV-2 in nasopharyngeal samples to overcome the limitations of virus isolation using conventional cell culture techniques. To do so, we developed an in-house flow cytometry (FC)-based surrogate assay (FlowSA) to detect the SARS-CoV-2 nucleocapsid (N) protein in Vero-E6 cells exposed to either pure laboratory-cultured viruses or those present in RT-PCR positive nasopharyngeal specimens from individuals participating in a prospective longitudinal cohort study from the time of infection (13). Our results indicate that, FlowSA can detect infectious viruses in clinical samples with Ct values as high as 39.

## Methods

### Cell culture

Vero E6 cells (ATCC CRL-1586) were thawed and cultured in Dulbecco’s minimal essential medium (DMEM) (Gibco, USA) supplemented with 10% fetal bovine serum (FBS) (v/v) (Life Science Production, UK), 1% penicillin (100 U/mL), and 1% streptomycin (100 U/mL) (v/v) (Gibco, USA).

### Production and Characterization of SARS-CoV-2 parent strain & variants

The reference SARS-CoV-2 strain (D614G), Alpha variant (B.1.1.7), and Delta variant (B.1.617.2) were obtained from the European Virus Archive global (EVAg-010V-03903) or from patients who presented at the diagnostic lab at University Medical Center Groningen. The patient-derived viruses were filtered through 0.22μm filter and isolated by passaging twice in Vero E6 cells cultured in DMEM supplemented with 2% FBS (v/v) and antibiotics. After second passage, the virus supernatants were harvested, snap-frozen, and stored at −80°C. Infectious SARS-CoV-2 viral titers were determined using a plaque assay, as described previously (14).

### SARS-CoV-2 RT-PCR

Viral load kinetics and detection of SARS-CoV-2 RNA in specimens (nasopharyngeal swabs) from the cohort of non-hospitalized COVID-19 patients were determined using an in-house protocol (6). The PCR assay was directed towards the SARS-CoV-2 E gene. Data analysis was performed using FlowG middleware software (LabHelp Labautomation). Samples with a Ct value lower than 34 were considered positive, while Ct values in the range of 34-39 were considered inconclusive, and the sample was repeated. Finally, Ct values greater than 40 were considered negative.

### FlowSA and CPE assay

The flowcytometry-based surrogate assay (FlowSA) was developed as a faster and more sensitive tool for assessing viral infectivity than the CPE assay (Figure 1).

**Figure 1.**
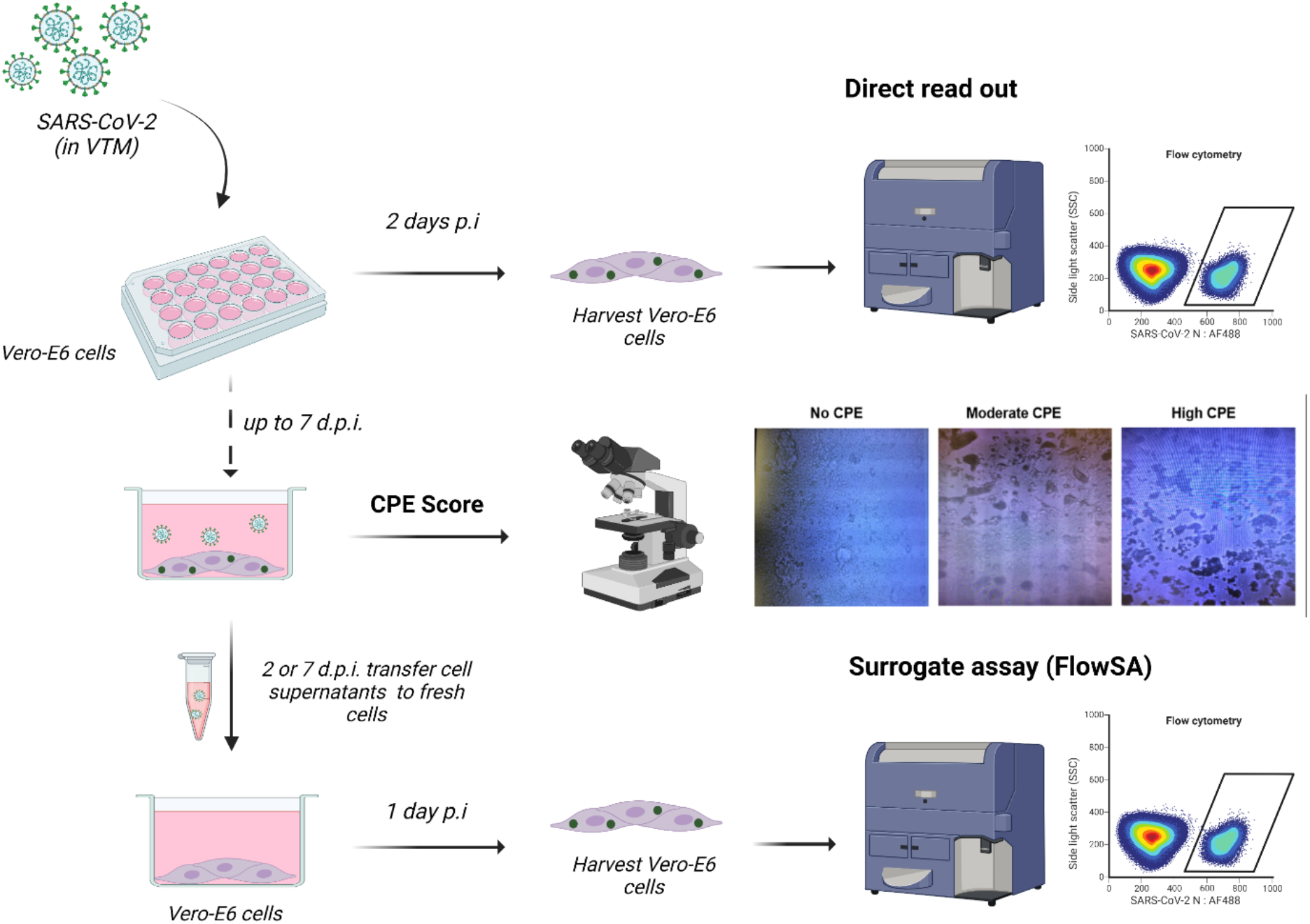
Schematic overview of direct infection assay, flow cytometry-based surrogate assay (FlowSA) and CPE-based approaches to detect infectious SARS-CoV-2 which have an experimentation time of 2, 3 and 7 days respectively. Abbreviations: CPE, cytopathic effect; d.p.i., day(s) post-infection; SARS-CoV-2, severe acute respiratory syndrome coronavirus 2; VTM, virus transport medium. Created with BioRender.

#### I. Infection

Briefly, 24-well plates were seeded with 1 × 10^4^ Vero-E6 cells and incubated for 24 h. The cells were then incubated with 200ul of serially diluted SARS-CoV-2, starting at a concentration of 7 × 10^5^ PFU/ml. After 2 hours, 300µL DMEM supplemented with 2% FBS was added to the wells and incubated at 37°C and 5% CO_2_ for 7 days. Each sample was tested in parallel in three separate wells: one for CPE assay and the other two wells for FlowSA.

#### II. CPE assay

The wells designated for CPE assay were observed daily under the microscope for a period of 7 days after incubation with virus. Cell morphology and the day of appearance of CPE was recorded.

#### III. Surrogate assay

On days 2 and 7 after infection, the supernatants were harvested and added to a new well of a 24-well plate seeded with 1×10^4^ Vero-E6 cells. The incubation was done for 12-14 hours. Cells from the respective wells were harvested by incubation with 1x trypsin and resuspended in FACS buffer [1x phosphate buffered saline (PBS) with 3% fetal bovine serum (FBS)]. This was followed by fixation with 4% paraformaldehyde and permeabilization with a permeabilization buffer (FACS buffer with 1% Tween-20). The cells were then stained with rabbit anti-SARS-CoV-2 N antibody (Invitrogen, Dilution 1:300), followed by staining with chicken anti-rabbit secondary antibody conjugated with Alexa Fluor 647 (AF647) (Invitrogen, Dilution 1:1000). Samples were acquired using a BD FACSAria cytometer and analyzed using Kaluza (Beckman Coulter) or FlowJo (BD Biosciences) flow cytometry analysis software.

### Clinical samples

Nasopharyngeal swabs were obtained from a cohort of non-hospitalized individuals who tested positive for COVID-19 as part of a longitudinal observational study which was initiated at the beginning of the COVID-19 pandemic in the Netherlands. The study design protocol of this cohort including ethics information and documentation has been described elsewhere (13). Briefly, this study has been approved by the Medical Ethical Review Committee of the UMCG (METc 2020/158) and follows international standards for the ethical conduct of research involving human subjects. All procedures employed in the clinical and related laboratory studies comply with national and European legislation in respect of research involving human subjects.

## Results

### Surrogate assay increases sensitivity of the infectious virus detection in virus samples with high Ct values

To evaluate the effectiveness of FlowSA in detecting low levels of infectious virus compared to conventional CPE or direct infection measurement by flow cytometry, we conducted a comparative analysis, as illustrated in Figure 1. To identify SARS-CoV-2 infection, we validated a previously established detection method for the viral N protein (10,15,16) (Supplementary Figure 1). Next, we used passage 2 of purified clinical isolate SARS-CoV-2 (D614G) circulating at that time in the Netherlands (14). Accordingly, 10-fold dilutions of the infectious virus ranging from 7×10^5^ to 7×10^−6^ PFU/mL (detected by qPCR between Ct values ranging from to 13-45) were prepared in virus transport medium (VMT) and tested with our in-house PCR assay for SARS-CoV-2 E gene (6). As expected, we observed an inverse correlation between virus dilutions and their corresponding Ct values (Supplementary Figure 2). Serial dilutions of SARS-CoV-2 were used to infect Vero-E6 cells, as shown in Figure 1. Cells seeded for the CPE assay were graded for the magnitude of CPE based on visual observation for seven days. In the CPE-based assay, we observed that the day of CPE appearance was inversely correlated with the concentration of infectious virus in VMT (Table 1). Notably, viral dilutions with high Ct values (>30) for SARS-CoV-2 PCR showed no CPE throughout the duration of the assay (Table 1).

**Table 1.**
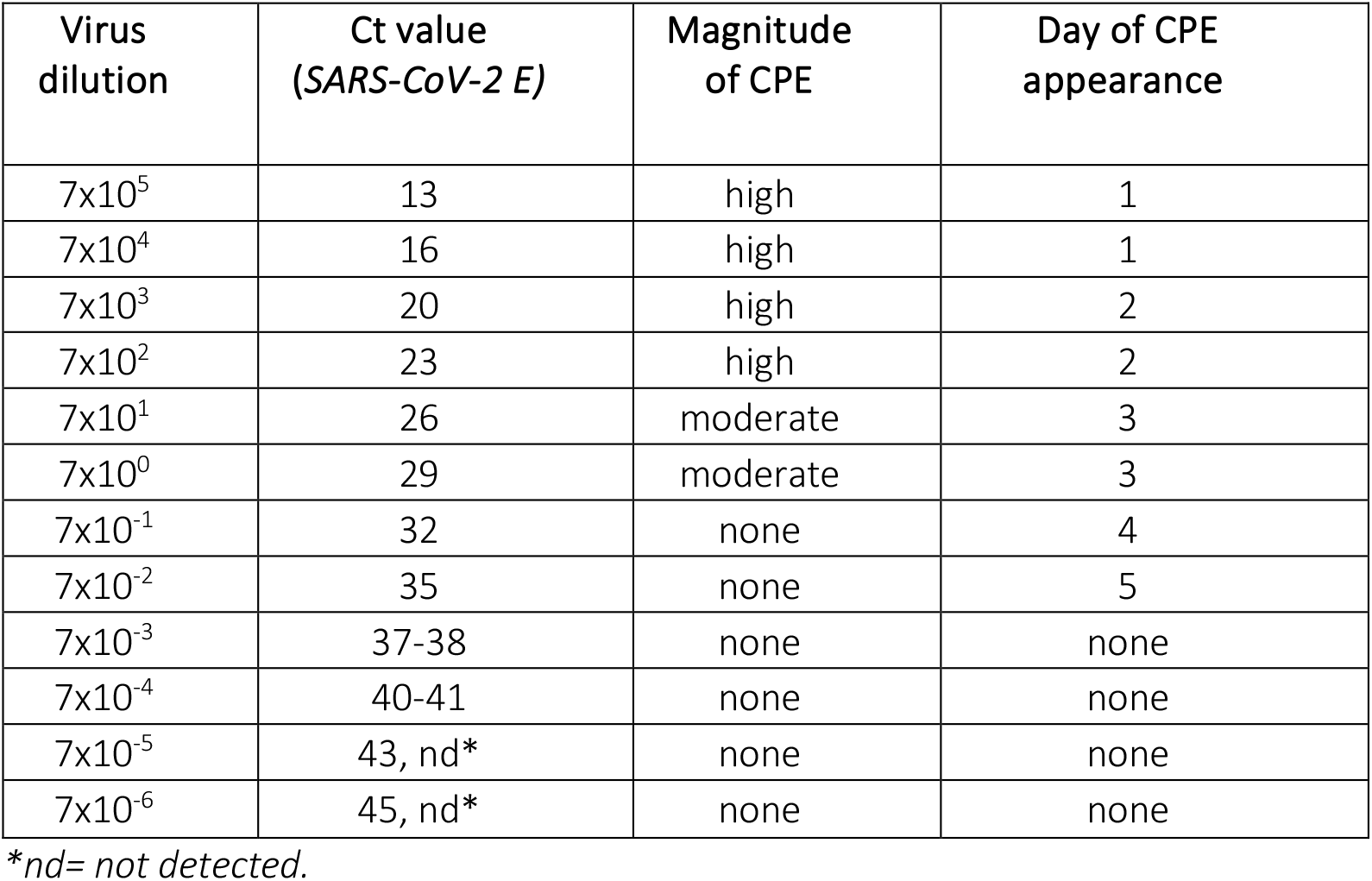
The serially diluted SARS-CoV-2 D614G strain was incubated on Vero-E6 cells and assessed for the appearance of CPE. Columns indicate the corresponding Ct values of serially diluted samples for SARS-CoV-2 PCR. The magnitude of CPE was determined based on the visual observation of each well and classified as high, moderate, or none (as shown in Figure 1).

**Figure 2.**
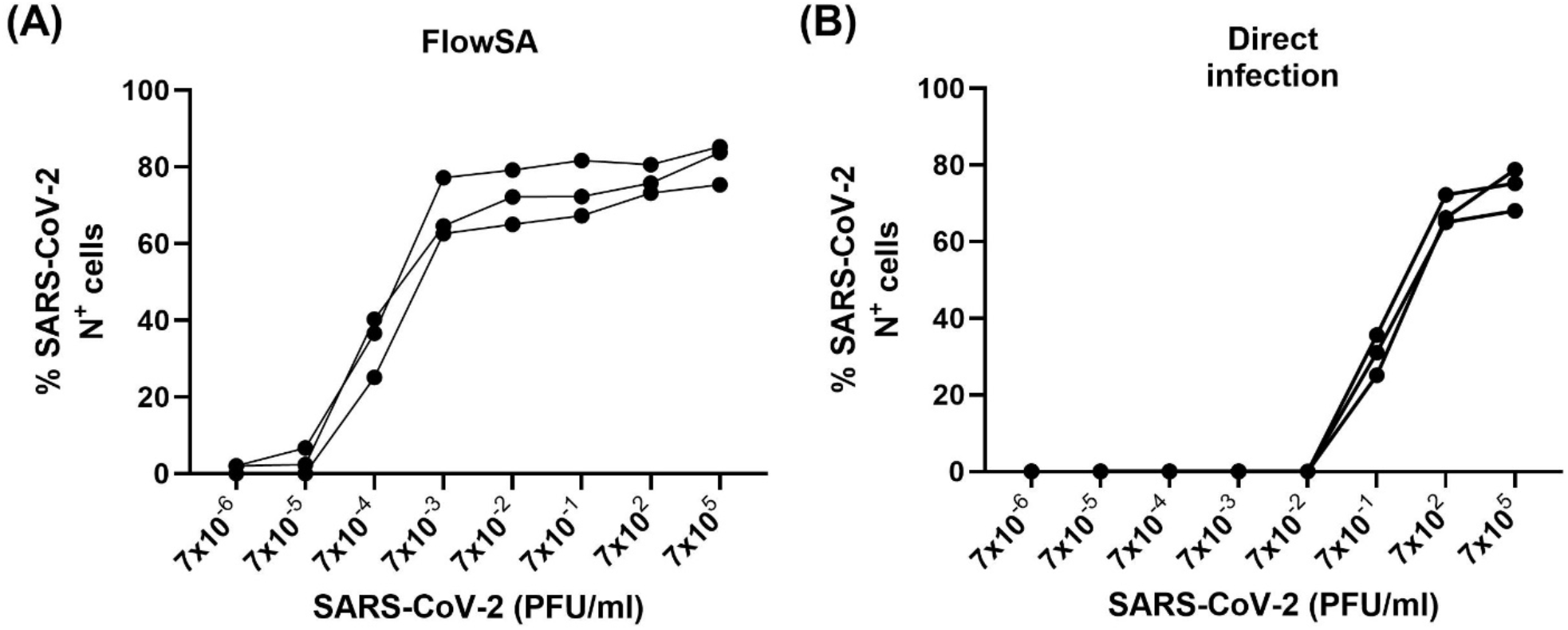
Comparison of SARS-CoV-2 infection measured using (A) FlowSA and (B) direct infection assay; both 48h post-infection, as shown in Figure 1. The data presented are from three independent experiments.

In parallel, to compare the direct infection assay with FlowSA, the serially diluted SARS-CoV-2 D614G strain was incubated with Vero-E6 cells in duplicate in 24-well plates (Figure 1). For the direct infection assay, cells harvested at 48 hours post-infection (hpi) were subjected to cytometry. For FlowSA, the supernatant was harvested and used to infect a fresh batch of Vero-E6 cells, which were analyzed by flow cytometry at 12-14 hpi. Here, we observed that high concentrations of the virus (7×10^2^ and 7×10^5^ PFU/ml) resulted in similar percentages of infected cells (Figure 2). However, in the surrogate assay, infected cells could be detected in wells treated with supernatants from cells infected with very low concentrations of the virus (∼7×10^−4^ PFU/ml), suggesting that the additional culturing step in the surrogate assay improves the limit of detection and may allow for the detection of viable SARS-CoV-2 at low titers in clinical samples.

### Validation of FlowSA for different SARS-CoV-2 variants

Next, we sought to ascertain the robustness of the surrogate assay in detecting various SARS-CoV-2 variants of public health concern that were circulating during the duration of the study. SARS-CoV-2 Alpha (B.1.1.7) or Delta (B.1.617.2) variants were independently isolated from qPCR-confirmed nasopharyngeal samples as per the protocol described in the Methods section, and serially diluted virus stocks were used to infect Vero-E6 cells, as described previously. Supernatants harvested on day 2 post-infection were used in the surrogate assay. The SARS-CoV-2 D614G strain was used as a reference strain for comparison. Indeed, we observed that FlowSA could detect infected cells with the Alpha and Delta variants (Supplementary Figure 3). Compared to the wild-type (D614G), the infectivity of the Alpha variant, although similar at the titer of 7×10^5^ PFU declined much faster and approached the detection limit already at 7×10^−1^ PFU/ml. The infectivity of the Delta variant, although slightly lower than that of the wild-type strain, resembled that of the wild-type strain and was detectable until a virus concentration of 7×10^−5^.

### Use of FlowSA for detection of infectious SARS-CoV-2 in clinical samples

Having established the ability of FlowSA to detect clinical isolates of multiple SARS-CoV-2 variants, we next tested samples obtained from an observational cohort of laboratory confirmed SARS-CoV-2 infected individuals (13). A total of 102 patient samples were analyzed using CPE and FlowSA assay systems. Approximately 39% (40/102) of the clinical samples were positive by the conventional CPE-based cell culture method (Figure 3A). Using FlowSA, approximately 80% (81/102) of the clinical samples were positive for SARS-CoV-2 N protein (Figure 3B). Remarkably, 42 samples that displayed no CPE in cell culture and thus would typically be deemed negative for viable SARS-CoV-2, were found positive using virus Ag-specific FlowSA (Figure 3C). In contrast, only 2 samples that showed CPE, were negative using FlowSA, suggesting that the CPE observed in these samples may not be induced by SARS-CoV-2. These samples had Ct values of 30 or more.

**Figure 3.**
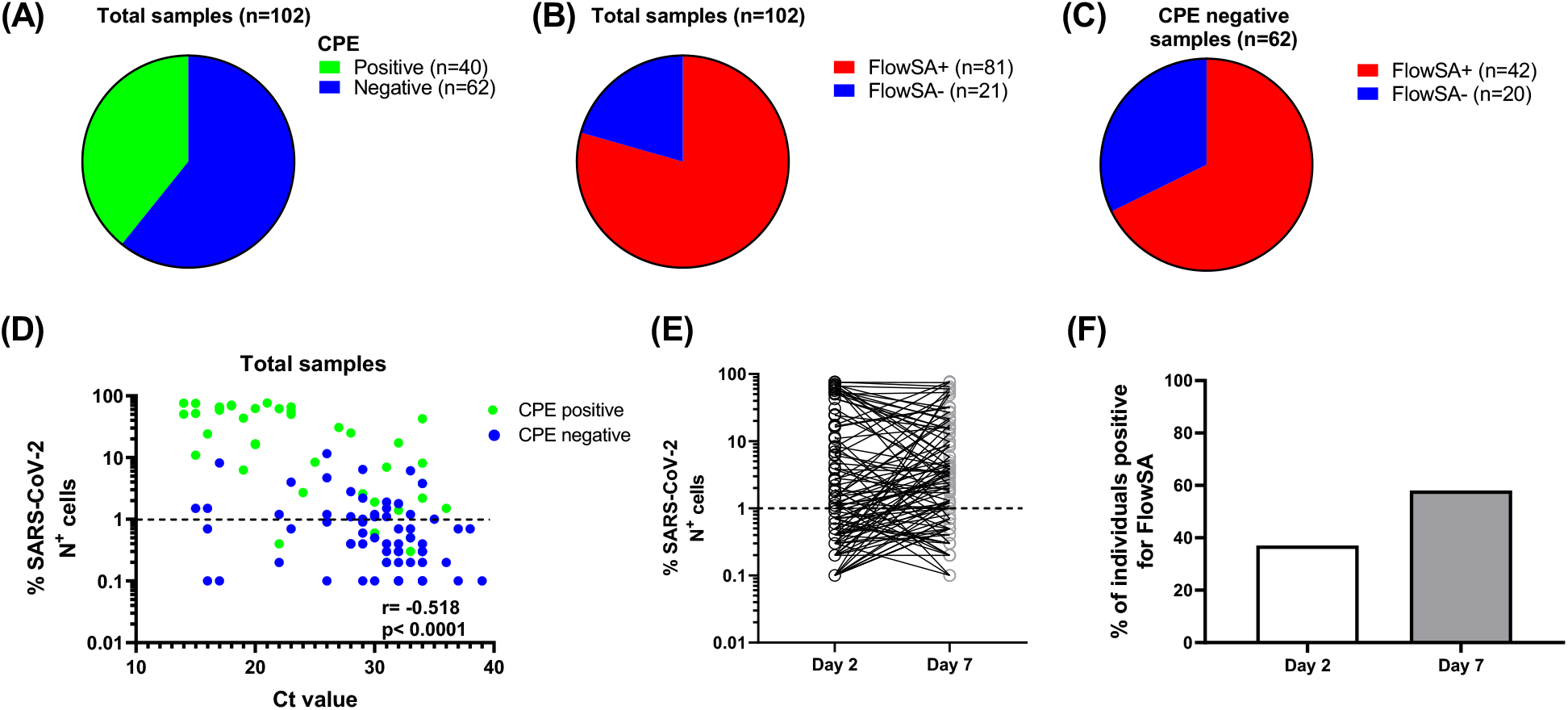
Analysis of nasopharyngeal swab samples from patients in the COVID-HOME study for SARS-CoV-2 using CPE method (A) and FlowSA (B). (C) Number of samples that were FlowSA+ and FlowSA– among samples that were negative for CPE-based method. (D) Comparison of FlowSA results from day 2 supernatants in COVID-Home patients and Ct values for the SARS-CoV-2 E gene in all patients. (E) Comparison of FlowSA results from day 2 and day 7 supernatants by CPE-based method (positive/negative). (F) Percentage of individuals positive for FlowSA on days 2 and 7 of testing.

The surrogate assay was also useful for detecting live viruses in supernatants obtained on day 2 from patient samples with varying Ct values in the SARS-CoV-2 PCR, irrespective of whether they showed CPE in Vero-E6 cells (Figure 3D).

Next, we sought to understand whether increasing the incubation time in Vero cells would further improve the sensitivity of FlowSA in comparison to CPE. To do so, we compared the FlowSA scores of cell supernatants collected on day 2 with those collected on day 7 (Figure 3E). Majority of the samples were positive for FlowSA on both day 2 and day 7. Interestingly, when we compared the FlowSA results of the 62 CPE negative samples, it was observed that performing FlowSA on day 7 led to 13 additional positive results (Figure 3F). Conversely, 9 samples which were positive for FlowSA on day 2 were negative on day 7.

In summary, FlowSA proves to be a useful tool for the detection of viable SARS-CoV-2 in clinical samples that are considered SARS-CoV-2 negative using conventional CPE-based methods.

## Discussion

The SARS-CoV-2 pandemic has highlighted the importance of developing a repertoire of sensitive and specific techniques for the detection of viruses in infected individuals. These techniques can complement routine diagnostic rapid tests and conventional RT-PCR and provide clinicians and public health officials with further insight into viral replication dynamics in infected individuals and influence decisions on community health.

In this study, we optimized a flow cytometry-based surrogate assay for the detection of the SARS-CoV-2 N protein in infected Vero-E6 cells. This simple and sensitive technique efficiently detected the highly conserved viral N protein from all the SARS-CoV-2 variants, including Alpha and Delta variants, which were prevalent during the course of the study.

Interestingly, the sensitivity of the assay was not equal across the variants analyzed, suggesting that infectivity and viral replication kinetics change as the virus evolves. Although we were unable to validate FlowSA with the currently circulating Omicron variants, the objective of this study was to develop a prototype assay on a proof of principle basis, which could be rapidly modified in response to SARS-CoV-2 variants as they emerge. Recently, Vanhulle et al. successfully stained cells infected with a reference Omicron variant using a SARS-CoV-2 N antibody, implying that the assay may also be applicable to the current strains (12). Further confirmation of this would be necessary in clinical samples.

The ultimate marker for transmission potential of SARS-CoV-2 has yet to be found, keeping in mind the interplay between viral and non-viral factors (17). During the pandemic we learned that SARS-CoV-2 RNA shedding, detected by PCR, can be prolonged and is probably not specific enough to be a marker for transmissibility (10). Another method for virus detection, the lateral flow assay, is not as sensitive as PCR and by that is thought to be suitable for the identification of infectious individuals (18). Current cell culture-based techniques used for the detection of viable viruses in clinical samples are cumbersome and depend on the development of CPE in infected cells, by which issues of sensitivity and specificity may occur. Using FlowSA, we were able to detect SARS-CoV-2 infection in cells infected with a viral input of less than 7×10^−1^ PFU/ml. Furthermore, we detected cells infected with SARS-CoV-2 in clinical samples that were culture-negative. Whether this technical sensitivity translates into clinical significance for transmission potential has to be established.

Owing to the simplicity of the surrogate assay, FlowSA may also be standardized to form a high-throughput assay and deployed for the testing of large numbers of clinical samples, as seen during surges or waves in the transmission of SARS-CoV-2, and to determine the patterns of shedding of viable virus in these individuals. Thus, this method may prove to be a useful tool for viral immunology assays.

## Data Availability

All data produced in the present work are contained in the manuscript

https://www.medrxiv.org/content/10.1101/2022.08.14.22278762v1.full

https://journals.plos.org/plosone/article?id=10.1371/journal.pone.0273599

## References

1. WHO. 11 March 2020. Coronavirus disease (COVID-19) situation report 51. https://www.who.int/docs/default-source/coronaviruse/situation-reports/20200311-sitrep-51-covid-19.pdf?sfvrsn=1ba62e57810

2. Wu F, Zhao S, Yu B et al. A new coronavirus associated with human respiratory disease in China. Nature 579(7798), 265–269 (2020).

3. Harvey, W.T., Carabelli, A.M., Jackson, B. et al. SARS-CoV-2 variants, spike mutations and immune escape. Nat Rev Microbiol 19, 409–424 (2021). 10.1038/s41579-021-00573-0

4. Bruce EA, Mills MG, Sampoleo R, Perchetti GA, Huang ML, Despres HW, Schmidt MM, Roychoudhury P, Shirley DJ, Jerome KR, Greninger AL, Botten JW. Predicting infectivity: comparing four PCR-based assays to detect culturable SARS-CoV-2 in clinical samples. EMBO Mol Med. 2022 Feb 7;14(2):e15290. doi: 10.15252/emmm.202115290.

5. Wang W, Xu Y, Gao R, et al. Detection of SARS-CoV-2 in Different Types of Clinical Specimens. JAMA. 2020;323(18):1843–1844. doi:10.1001/jama.2020.3786

6. Gard L, Fliss MA, Bosma F, Ter Veen D, Niesters HGM. Validation and verification of the GeneFinder™ COVID-19 Plus RealAmp kit on the ELITe InGenius® instrument. J Virol Methods. 2022 Feb;300:114378. doi: 10.1016/j.jviromet.2021.114378.

7. Laferl H, Kelani H, Seitz T, Holzer B, Zimpernik I, Steinrigl A, Schmoll F, Wenisch C, Allerberger F. An approach to lifting self-isolation for health care workers with prolonged shedding of SARS-CoV-2 RNA. Infection. 2021 Feb;49(1):95–101. doi: 10.1007/s15010-020-01530-4.

8. Funk, D.J., Bullard, J., Lother, S. et al. Persistence of live virus in critically ill patients infected with SARS-COV-2: a prospective observational study. Crit Care 26, 10 (2022). 10.1186/s13054-021-03884-z

9. Kim DY, Lin MY, Jennings C, Li H, Jung JH, Moore NM, Ghinai I, Black SR, Zaccaro DJ, Brofman J, Hayden MK; CDC Prevention Epicenter Program. Duration of replication-competent SARS-CoV-2 shedding among patients with severe or critical coronavirus disease 2019 (COVID-19). Clin Infect Dis.

10. van Kampen JJA, van de Vijver DAMC, Fraaij PLA, Haagmans BL, Lamers MM, Okba N, van den Akker JPC, Endeman H, Gommers DAMPJ, Cornelissen JJ, Hoek RAS, van der Eerden MM, Hesselink DA, Metselaar HJ, Verbon A, de Steenwinkel JEM, Aron GI, van Gorp ECM, van Boheemen S, Voermans JC, Boucher CAB, Molenkamp R, Koopmans MPG, Geurtsvankessel C, van der Eijk AA. Duration and key determinants of infectious virus shedding in hospitalized patients with coronavirus disease-2019 (COVID-19). Nat Commun. 2021 Jan 11;12(1):267. doi: 10.1038/s41467-020-20568-4.

11. Hiroi S, Kubota-Koketsu R, Sasaki T, Morikawa S, Motomura K, Nakayama EE, Okuno Y, Shioda T. Infectivity assay for detection of SARS-CoV-2 in samples from patients with COVID-19. J Med Virol. 2021 Oct;93(10):5917–5923. doi: 10.1002/jmv.27145.

12. Vanhulle E, Provinciael B, Stroobants J, Camps A, Maes P, Vermeire K. Intracellular flow cytometry complements RT-qPCR detection of circulating SARS-CoV-2 variants of concern. Biotechniques. 2022 Jun;72(6):245–254. doi: 10.2144/btn-2022-0018. 2022 May 24:ciac405. doi: 10.1093/cid/ciac405.

13. Tami A, van der Gun BTF, Wold KI, Vincenti-González MF, Veloo ACM, Knoester M, Harmsma VPR, de Boer GC, Huckriede ALW, Pantano D, Gard L, Rodenhuis-Zybert IA, Upasani V, Smit J, Dijkstra AE, de Haan JJ, van Elst JM, van den Boogaard J, O’ Boyle S, Nacul L, Niesters HGM, Friedrich AW. The COVID HOME study research protocol: A prospective cohort study of non-hospitalized COVID-19 patients. PLoS One. 2022 Nov 3;17(11):e0273599. doi: 10.1371/journal.pone.0273599.

14. Ter Ellen BM, Dinesh Kumar N, Bouma EM, Troost B, van de Pol DPI, van der Ende-Metselaar HH, Apperloo L, van Gosliga D, van den Berge M, Nawijn MC, van der Voort PHJ, Moser J, Rodenhuis-Zybert IA, Smit JM. Resveratrol and Pterostilbene Inhibit SARS-CoV-2 Replication in Air-Liquid Interface Cultured Human Primary Bronchial Epithelial Cells. Viruses. 2021 Jul 10;13(7):1335. doi: 10.3390/v13071335

15. Cubuk J, Alston JJ, Incicco JJ, Singh S, Stuchell-Brereton MD, Ward MD, Zimmerman MI, Vithani N, Griffith D, Wagoner JA, Bowman GR, Hall KB, Soranno A, Holehouse AS. The SARS-CoV-2 nucleocapsid protein is dynamic, disordered, and phase separates with RNA. Nat Commun. 2021 Mar 29;12(1):1936. doi: 10.1038/s41467-021-21953-3

16. Bai Z, Cao Y, Liu W, Li J. The SARS-CoV-2 nucleocapsid protein and its role in viral structure, biological functions, and a potential target for drug or vaccine mitigation. Viruses 13(6), e13061115 (2021).

17. Wang CC, Prather KA, Sznitman J, Jimenez JL, Lakdawala SS, Tufekci Z, Marr LC. Airborne transmission of respiratory viruses. Science. 2021 Aug 27;373(6558):eabd9149. doi: 10.1126/science.abd9149.

18. Deeks JJ, Singanayagam A, Houston H, Sitch AJ, Hakki S, Dunning J, Lalvani A. SARS-CoV-2 antigen lateral flow tests for detecting infectious people: linked data analysis. BMJ. 2022 Feb 23;376:e066871. doi: 10.1136/bmj-2021-066871.

19. Demuth S, Damaschek S, Schildgen O, Schildgen V. Low sensitivity of SARS-CoV-2 rapid antigen self-tests under laboratory conditions. New Microbes New Infect. 2021 Sep;43:100916. doi: 10.1016/j.nmni.2021.

