## Supplementart Figure 1 for "Evaluation of a flow cytometry-based surrogate assay (FlowSA) for the detection of SARS-CoV-2 in clinical samples"

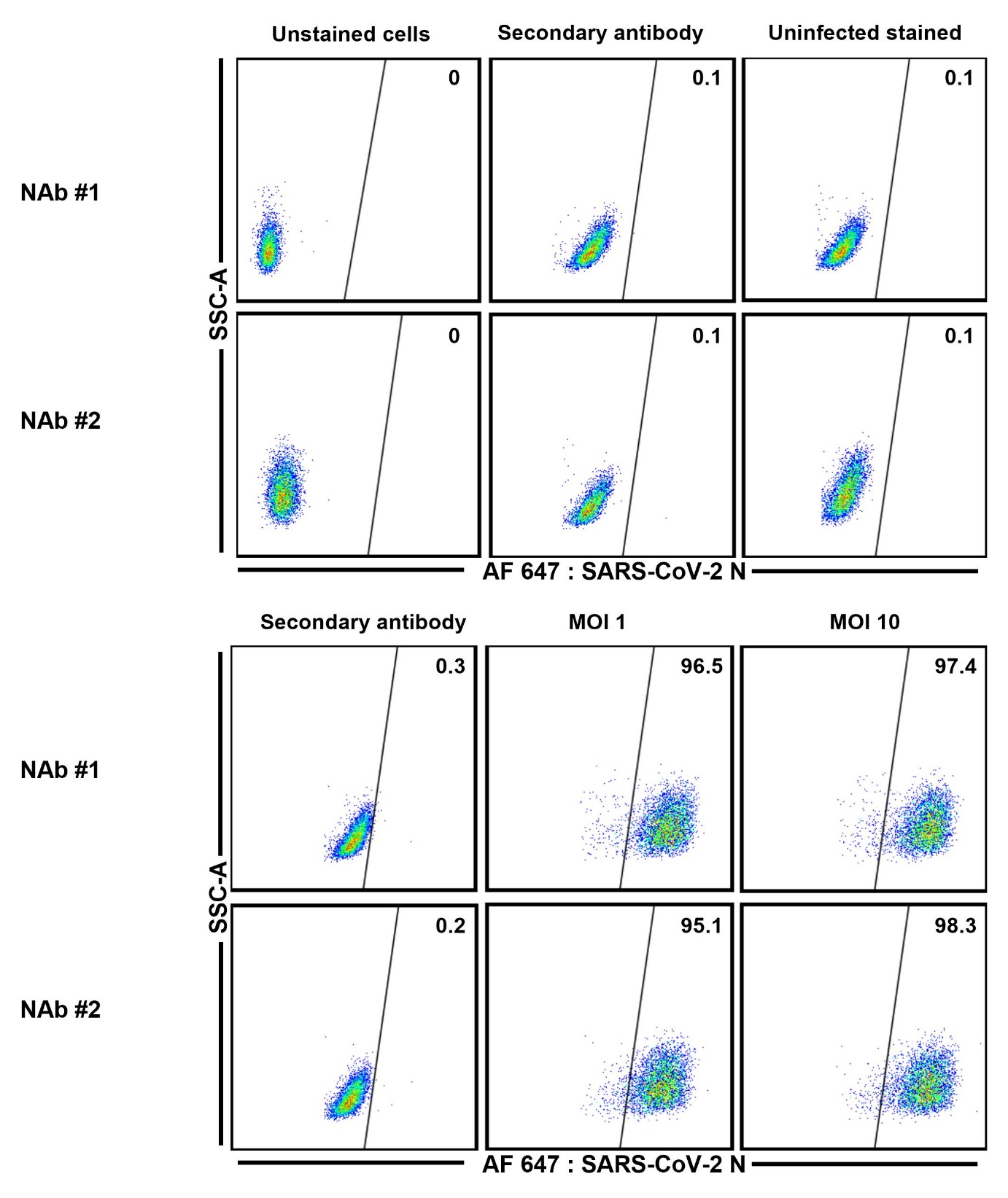

**Supplementary Figure 1:** Validation of SARS-CoV-2 infection detection using two different lots of anti-nucleocapsid (N) antibody (NAb #1 & NAb #2). To detect SARS-CoV-2 infection, Vero-E6 cells were infected with the SARS-CoV-2/NL/2020 strain at MOI (multiplicity of infection) of 1 and 10 for 24 h and stained with SARS-CoV-2 N antibody. As shown in the scheme, stained uninfected Vero-E6 cells were used as negative controls for infection (Uninfected stained) and to determine background staining (Secondary antibody).
