## Supplementart Figure 2 for "Evaluation of a flow cytometry-based surrogate assay (FlowSA) for the detection of SARS-CoV-2 in clinical samples"

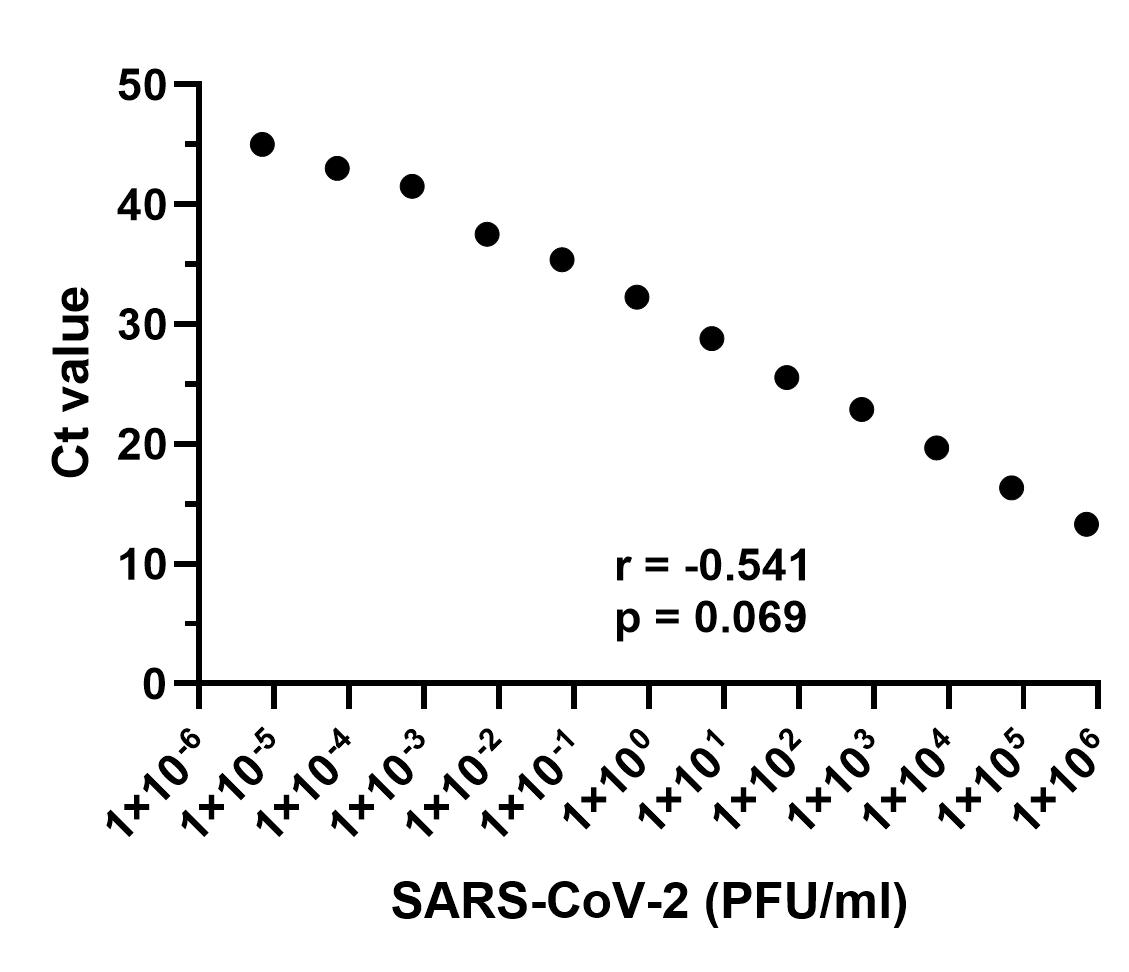

***Supplementary Figure 2.*** *Correlation between viral titer expressed as plaque forming units (PFU/ml) of SARS-CoV-2 and the corresponding Ct value observed in SARS-CoV-2 qRT-PCR.*
