## Supplementart Figure 3 for "Evaluation of a flow cytometry-based surrogate assay (FlowSA) for the detection of SARS-CoV-2 in clinical samples"

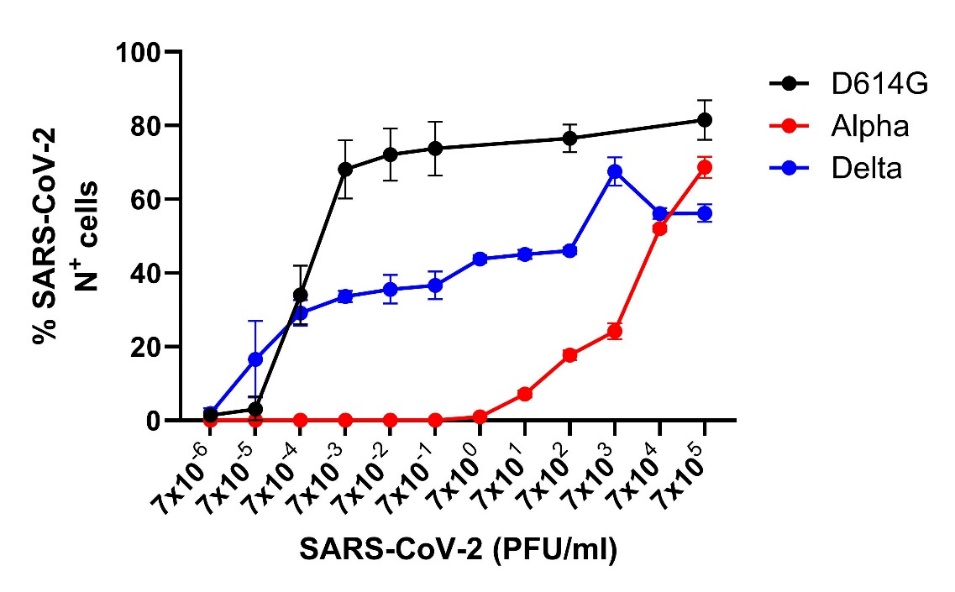

***Supplementary Figure 3:*** *Comparison of infectivity of various SARS-CoV-2 variants of concern –*D614G *strain, Alpha (B1.1.7), and Delta (B1.617.2)–using FlowSA. Data are presented as mean ± SEM of three independent experiments.*
